# Effects of temperature and humidity on the spread of COVID-19: A systematic review

**DOI:** 10.1101/2020.04.14.20064923

**Authors:** Paulo Mecenas, Renata Travassos da Rosa Moreira Bastos, Antonio Carlos Rosário Vallinoto, David Normando

## Abstract

**Background:** Faced with the global pandemic of COVID-19, declared by World Health Organization (WHO) on March 11^th^ 2020, and the need to better understand the seasonal behavior of the virus, our team conducted this systematic review to describe current knowledge about the emergence and replicability of the virus and its correlation with different weather factors such as temperature and relative humidity.

**Methods:** The review was registered with the PROSPERO database. The electronic databases PubMed, Scopus, Web of Science, Cochrane Library, LILACS, OpenGrey and Google Scholar were examined with the searches restricted to the years 2019 and 2020. Risk of bias assessment was performed using the Joanna Briggs Institute (JBI) Critical Appraisal Checklist tool. The GRADE tool was used to assess the quality of the evidence.

**Results:** The initial screening identified 517 articles. After examination of the full texts, seventeen studies met the review’s eligibility criteria. Great homogeneity was observed in the findings regarding the effect of temperature and humidity on the seasonal viability and transmissibility of COVID-19. Cold and dry conditions were potentiating factors on the spread of the virus. After quality assessment, four studies had a high risk of bias and thirteen studies were scored as moderate risk of bias. The certainty of evidence was graded as low for both outcomes evaluated.

**Conclusion:** Considering the existing scientific evidence, warm and wet climates seem to reduce the spread of COVID-19. The certainty of the evidence generated was graded as low. However, these variables alone could not explain most of the variability in disease transmission.

## Introduction

Respiratory tract infections are the most common infections worldwide, representing a source of significant morbidity and a considerable economic burden to health care.[1] The coronaviruses, *Orthocoronaviridae* sub-family, are so called for their crown-like spikes on the viral surface. They are classified into four main genus sub-groups known as *Alphacoronavius, Betacoronavirus, Gammacoronavirus, Deltacoronavirus*, and are able to infect human beings with a common flu.[2,3]

A new epidemic of Severe Acute Respiratory Syndrome (SARS) Coronavirus has emerged since December 2019, namely SARS-CoV-2 or COVID-19, in Wuhan, the capital of Hubei Province, China. An outbreak of atypical pneumonia named COVID-19 caused by this virus has been reported,[4,5] and the pattern of human-to-human transmissibility of the virus has occurred nationally and internationally.[6,7]

The etiological agents have been confirmed as a new subset of coronaviruses.[8] Spread of SARS-CoV-2, like other respiratory viruses, namely its predecessor SARS-CoV, can be due to easy aerial transmissions of respiratory droplets, exposing the virus to external environmental conditions.[9] This epidemic has caused a collapse on health care services and economies of affected countries, and the overall mortality rate was estimated to be 4.7%, but in elderly patients, aged 60 or above, it can increase to up 14.8%.[10] A notable feature of SARS-CoV-2 is its predilection for transmission in the health care setting and to close family and social contacts by different manners, such as droplets, close direct or indirect contact, but the relative importance of these routes of transmission is still unclear.[11] The transmission can be affected by a number of factors, including population density, migratory flow, host immunity, medical care quality and, presumably, climate conditions (such as temperature and humidity).[12,13]

Limited studies have investigated climate parameters as important factors that could influence the SARS-CoV-2 spread. The seasonal nature in the outbreaks of respiratory virus infections is a common phenomenon, with peaks often occurring in low temperatures, during the winter months.[1] The coronavirus can retain its infectivity up to 2 weeks in a low temperature and low humidity environment, which might facilitate the virus transmission in a community located in a subtropical climate.[11] The mechanism underlying these patterns of climate determination that lead to infection and possible disease transmission is associated with the ability of the virus to survive external environmental conditions before staying in a host.[9] Many etiological factors such as changes in host physiological susceptibility, immune system function, social behavior, and weather conditions have been suggested in this context.[14]

It is supposed that high temperature and humidity, together, have a combined effect on inactivation of coronaviruses while the opposite weather condition can support prolonged survival time of the virus on surfaces and facilitate the transmission and susceptibility of the viral agent.[11] This combination may trigger an impairment of the local and systemic antiviral defense mechanisms, leading to increased host susceptibility to the respiratory viruses in winter.[15]

Nevertheless, there is still divergence in the literature about the effects of temperature and humidity on the viability and transmissibility of the coronavirus infection that appeared in 2019. Faced with the global pandemic of COVID-19, declared by World Health Organization (WHO) on March 11^th^ 2020,[16] and the need to better understand the seasonal behavior of the virus, our team conducted this systematic review to describe current knowledge about the emergence and replicability of the virus and its correlation with different weather factors such as temperature and relative humidity. This information could be useful to develop and implement an efficient health information system with public interventions to control the incidence, and to curb the spread, of COVID-19 in the world.

## Methods

### 1.1. Protocol and registration

This systematic review was registered with the PROSPERO database (CRD42020176909), and performed according with the PRISMA (Preferred Reporting Items for Systematic Reviews and Meta-Analyses) guidelines (S1 file).[17]

### 1.2. Eligibility criteria

Manuscripts that evaluated the effects of different climatic conditions of temperature and/or humidity on the spread of COVID-19 were included. The search strategy was defined based on the PECOS format as follows:

> Population (P): Humans diagnosed with COVID-19;
>
> Exposition (E): Different weather conditions: humidity, temperature;
>
> Comparison (C): Without comparison;
>
> Outcome (O): spread of SARS-CoV-2 (COVID-19);
>
> Study design (S): Observational studies, prospective or retrospective, case reports, case-series.

The exclusion criteria involved studies that evaluated other upper and lower respiratory tract infections, such as Middle East Respiratory Syndrome Coronavirus (MERS-CoV), SARS-CoV and influenza. The assessment of other climatic conditions, except for temperature and humidity, was also not considered. Opinion articles, animal or laboratory studies, and literature reviews were not included.

### 1.3. Information sources

The following electronic databases were searched: PubMed, Scopus, Web of Science, Cochrane Library, LILACS, OpenGrey and Google Scholar. A hand search was also conducted by reading the references list of the articles included. No restriction on language has been applied. Date of publication has been limited to the year 2019 and 2020. The search was conducted up to March 24^th^, 2020 in all databases, and until 31^st^ March, 2020 only in Google Scholar.

### 1.4. Search Strategy and Study Selection

The electronic searches were performed independently by two authors (XX and XXX). In case of disagreements, a third author (XXXX) was consulted. The search strategy was developed through a combination of MeSHs, Entry Terms and Keywords related to the PECOS strategy using Boolean operators. The full search strategies for each database are illustrated in Table 1.

**Table 1.**
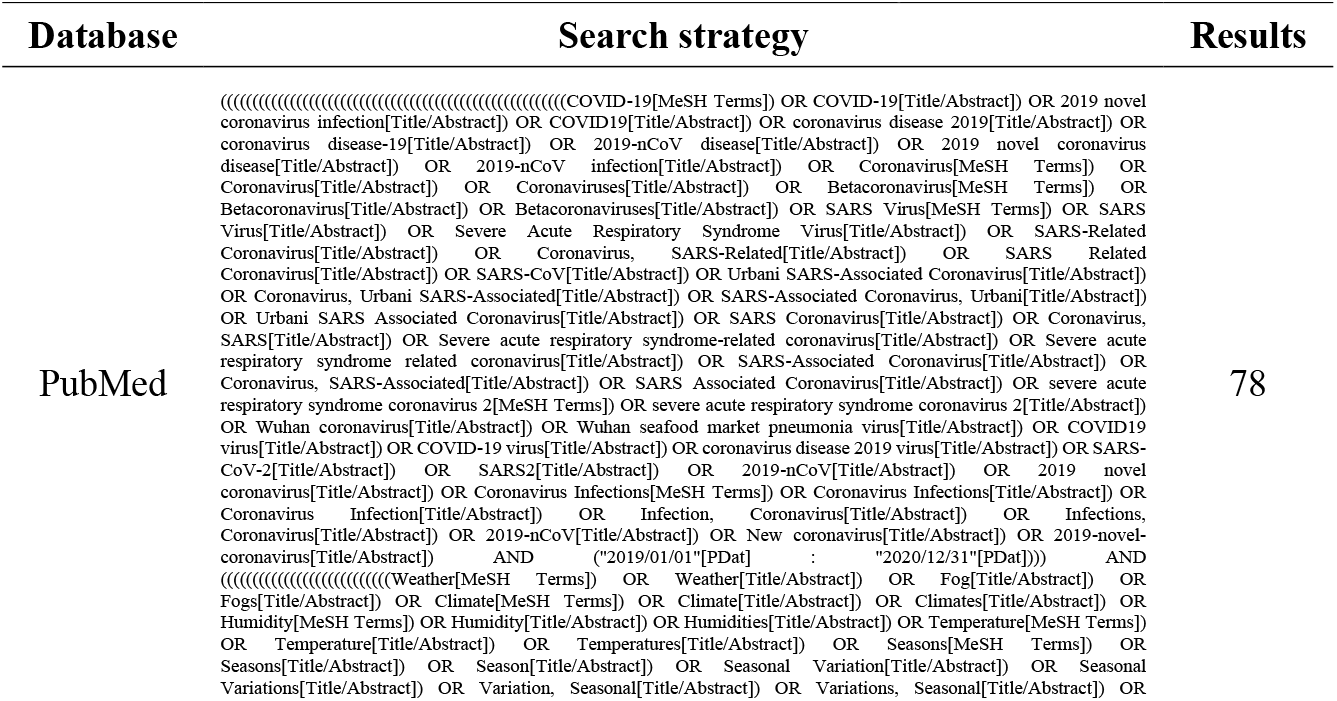
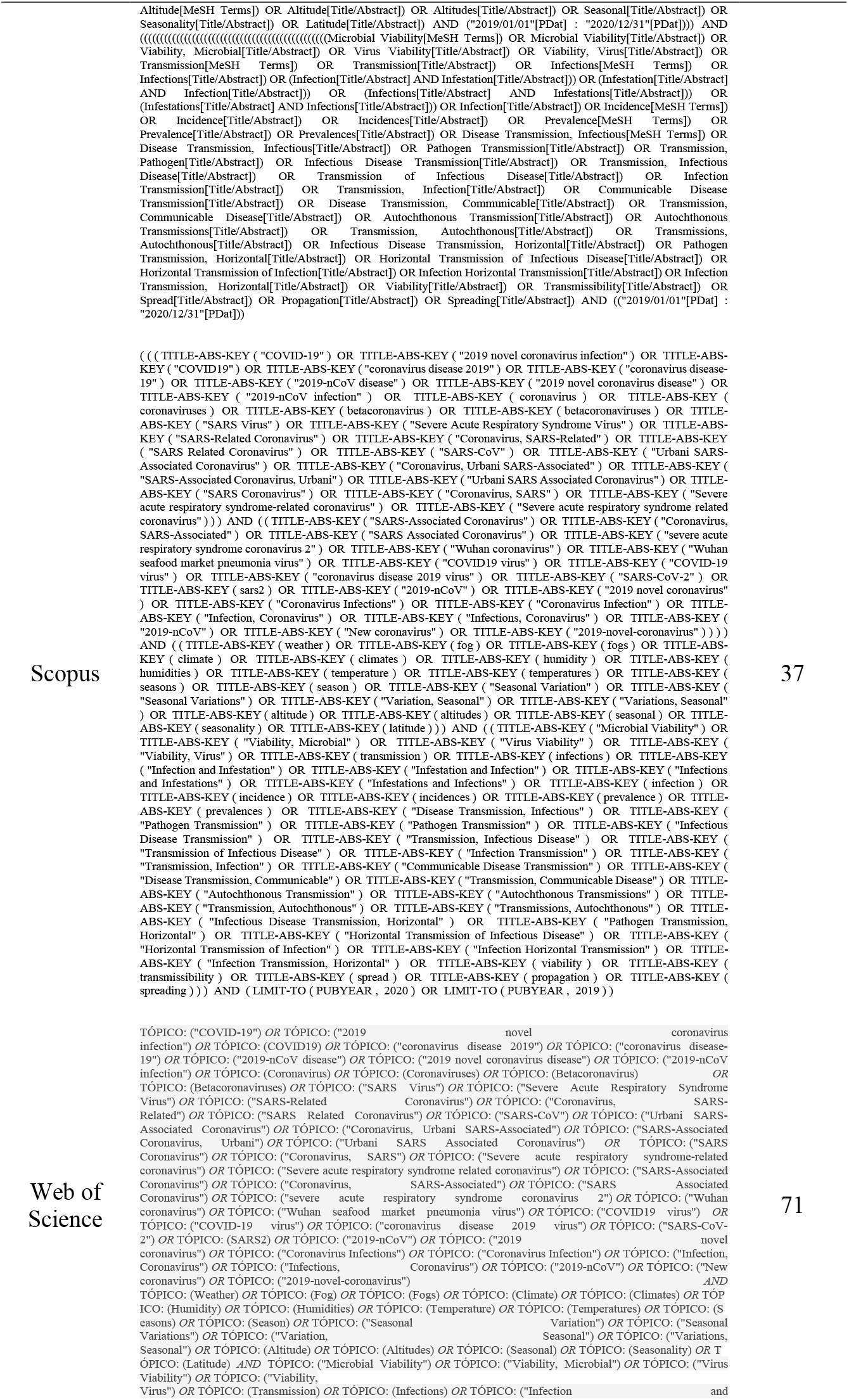
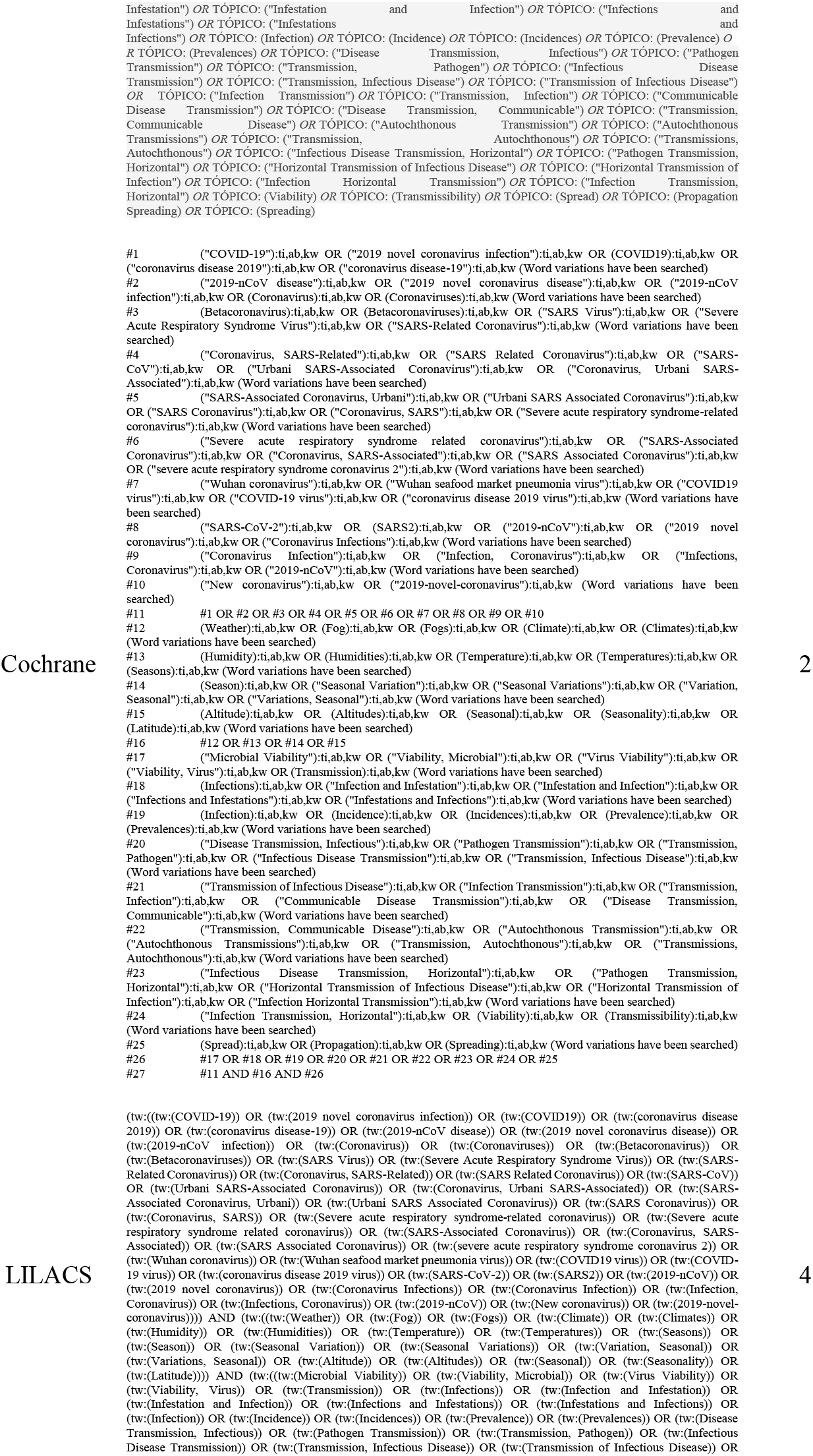
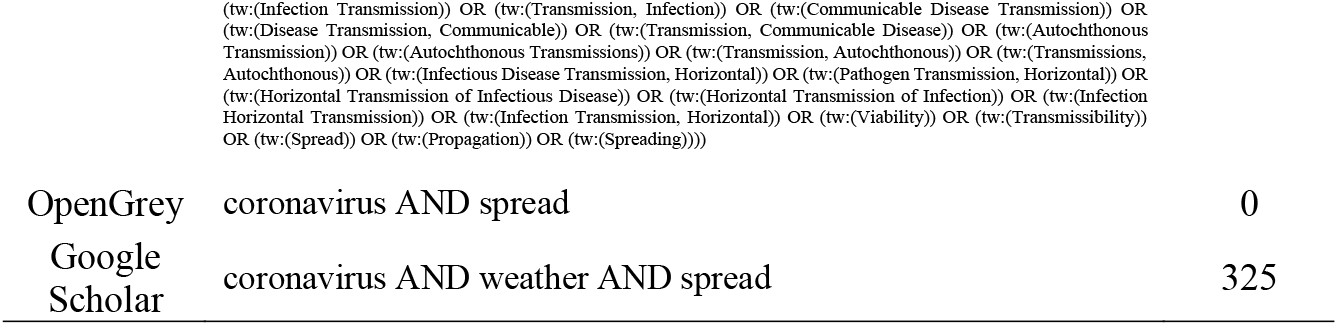
Search strategy for electronic databases.

The citations were saved in a reference manager (EndNote, x9 version, Clarivate Analytics, Philadelphia, PA, USA). After removing duplicates, titles and abstracts were analyzed according to the eligibility criteria. The selected articles were evaluated by full text, and a final selection was conducted.

### 1.5. Data extraction

Two authors collected the data independently (XX and XXX), extracting the following items: authors, year, location and type of study; date of COVID-19 data collection; date of meteorological data collection; sample countries; weather variables; COVID-19 data sources; meteorological data sources; statistical analysis and main results. Meta-analysis was planned if there was relative homogeneity of the data and the methods for obtaining it, for each selected article.

### 1.6. Assessment of risk of bias

All the included studies were assessed for methodological rigor using the Joanna Briggs Institute (JBI) Critical Appraisal Checklist tool.[18] The checklist for cross-sectional studies uses eight criteria. The evaluation content includes: the criteria for inclusion in the sample; the study subjects and the setting described; measurement of the exposure; the objective, standard criteria used for measurement of the condition; identifying the confounding factors; the strategies to deal with confounding factors; measurement of the outcomes; and the statistical analysis used. Each component was rated as “yes”, “no”, “unclear”, or “not applicable”. With 1-3 “yes” scores, the risk of bias classification is high, 4-6 “yes” scores are moderate and 7-8 are low risk of bias. The information about the studies was extracted, synthesized from the data, and reflected in the results and conclusions of this systematic review. Two authors (XX and XXX) independently evaluated the quality of each study, and disagreements were resolved by discussion within the review team.

### 1.7. Certainty of evidence

The included articles were given a narrative grade related to the outcomes assessed in this review (effects of temperature and humidity on spread of COVID-19) according to the GRADE tool (Grading of Recommendation, Assessment, Development, and Evaluation) (GRADEpro Guideline Development Tool, available online at gradepro.org).[19] This tool considers five aspects for rating the level of evidence: design, risk of bias, consistency, directness, and precision of the studies. The level of evidence was classified as high, moderate, low or very low. The outcomes evaluated were “Association between temperature and spread rate of COVID-19” and “Association between humidity and spread rate of COVID-19”.

## Results

### Study Selection

The initial searches identified 517 articles: 78 from PubMed, 37 from Scopus, 71 from Web of Science, 2 from Cochrane Library, 4 from LILACS, 0 from OpenGrey and 325 from Google Scholar. After exclusion of duplicates, 434 studies remained. After reading the titles and abstracts, 26 remaining articles were evaluated by full text and 9 were excluded. The reasons for exclusion are shown in Table 2. Seventeen studies were included and selected for qualitative analysis of risk of bias (Fig 1).

**Table 2.** List of excluded studies with reasons for exclusion.

| Reference | Reason for exclusion |
| --- | --- |
| Boulos et al. (2020) | Editorial article. |
| Cai et al. (2020) | The article evaluated the mortality rate of COVID-19. |
| Jackson et al. (2020) | The article did not evaluate COVID-19. |
| Khalifa et al. (2019) | The article did not evaluate COVID-19. |
| Ma et al. (2020) | The article evaluated daily mortality rate of COVID-19. |
| Moriyama et al. (2020) | Literature review. |
| Neher et al. (2020) | The article did not report temperature and humidity variables. |
| Rai et al. (2020) | Predictive study. |
| Zhao et al. (2020) | The article did not evaluate COVID-19. |

### Characteristics of included articles

The characteristics of the included studies are described in Table 3. All of them were retrospective observational studies that associated weather variables (temperature and humidity) with the spread of COVID-19.[9,20-35] Only two studies were also classified as prospective, as they both suggest future policies to prevent the spread of COVID-19 through additional control with a new vaccine,[25] and the use of the best fitted predictive model of the climatic conditions of the next 12 days in 5 cities worldwide.[24] Two papers investigated the effect of temperature only on seasonal variability in transmission of COVID-19.[21,30] Fifteen articles evaluated the effect of the variables under study - temperature associated with humidity - in transmission of SARS-CoV-2.[9,20,22-29,31-35] Of these, six[20,23-25,27,32] studies evaluated another constant variable, not included in this systematic review, that did not demonstrate an important factor if modeled alone in the transmission of the virus, the wind speed. It was not the objective of this systematic review to verify or discuss the statistical parameters in the manuscript. Therefore, we decided to perform a narrative synthesis, risk of bias and a narrative grade of evidence of the results.

**Table 3.**
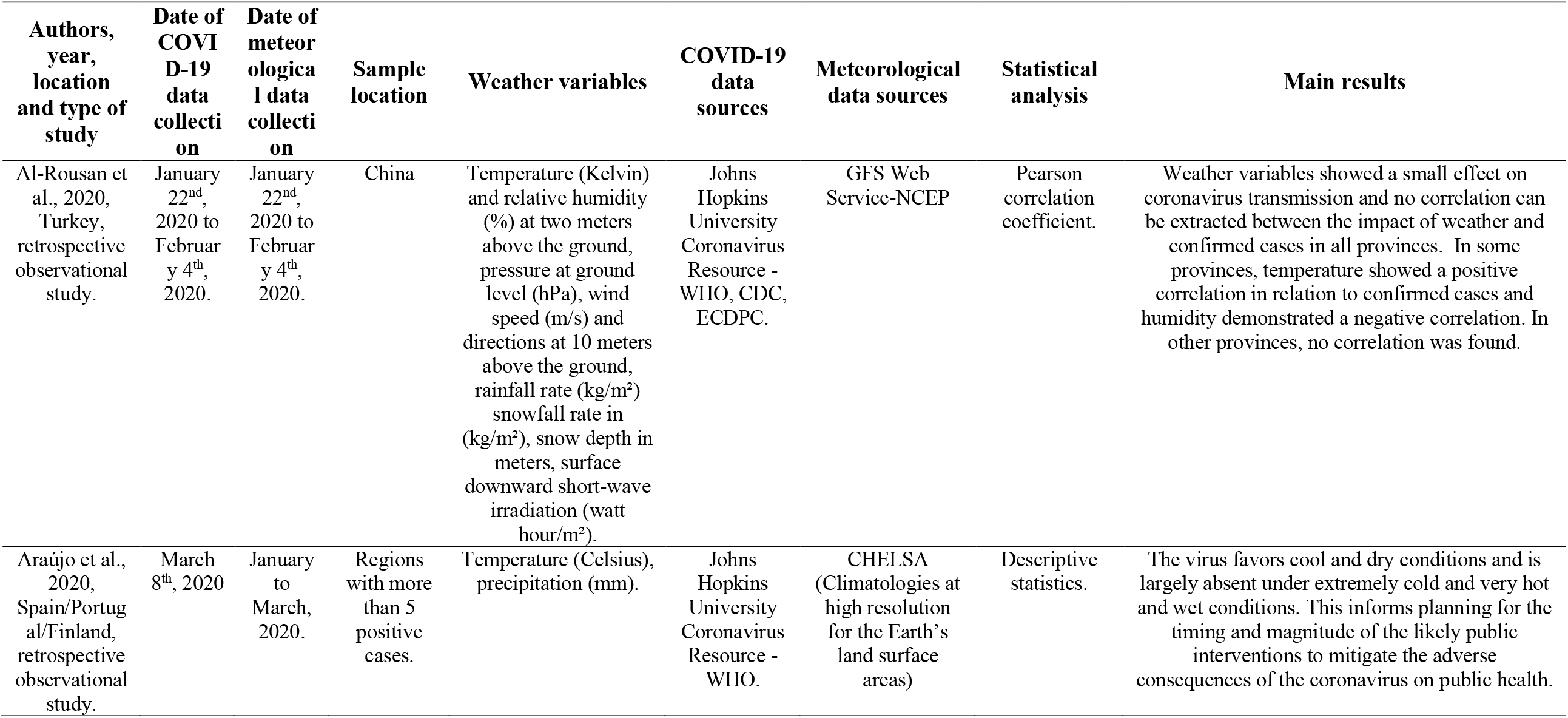
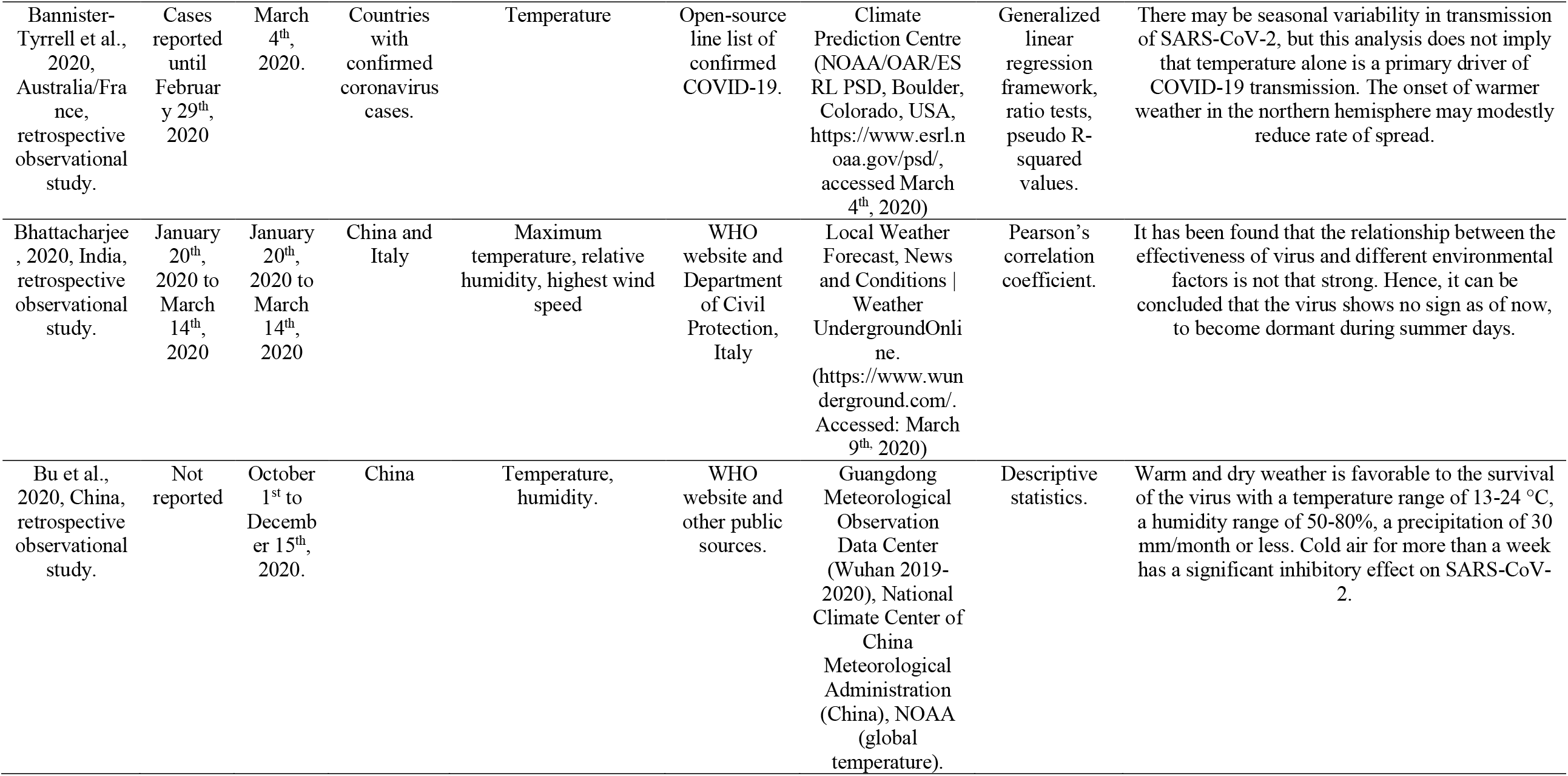
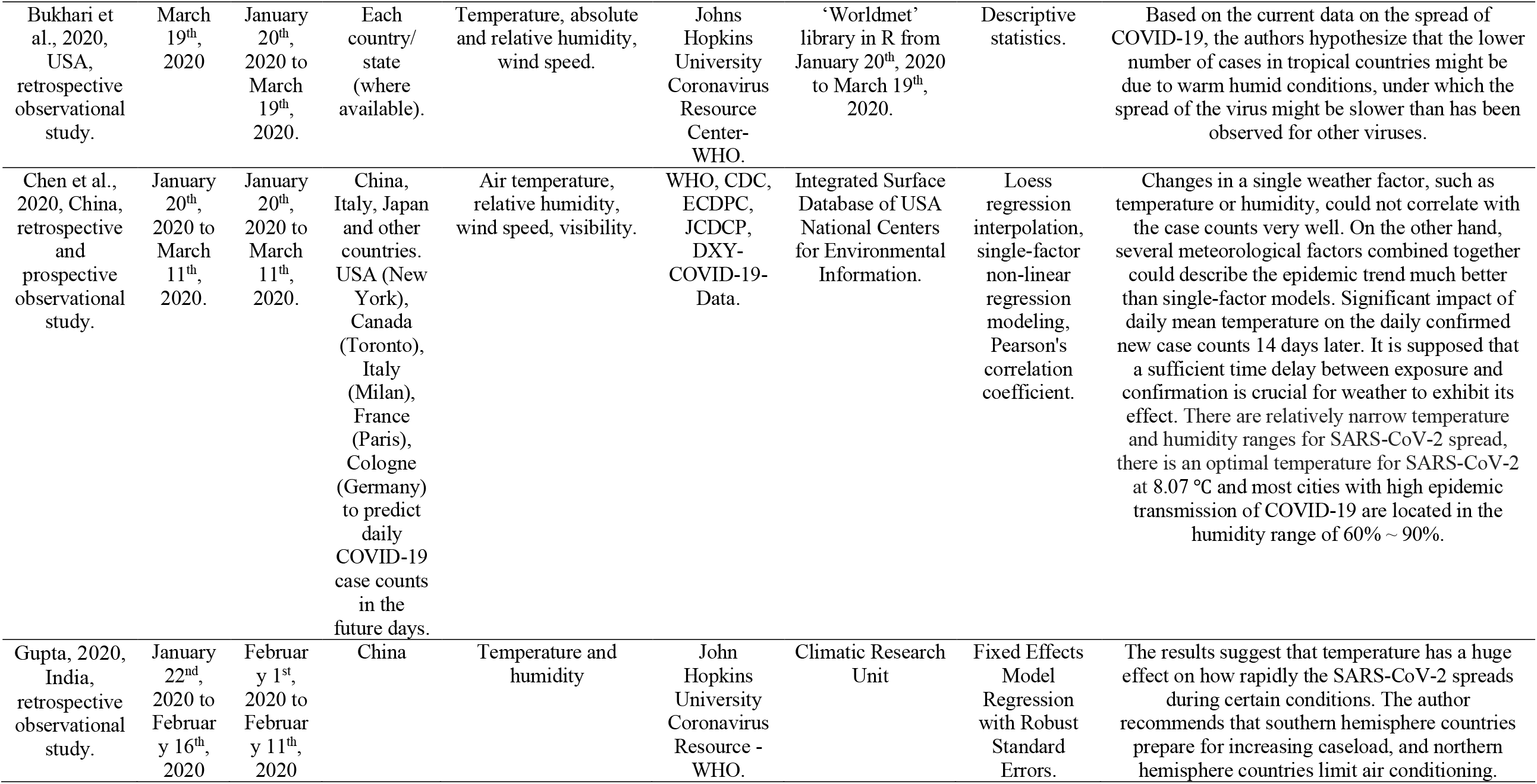
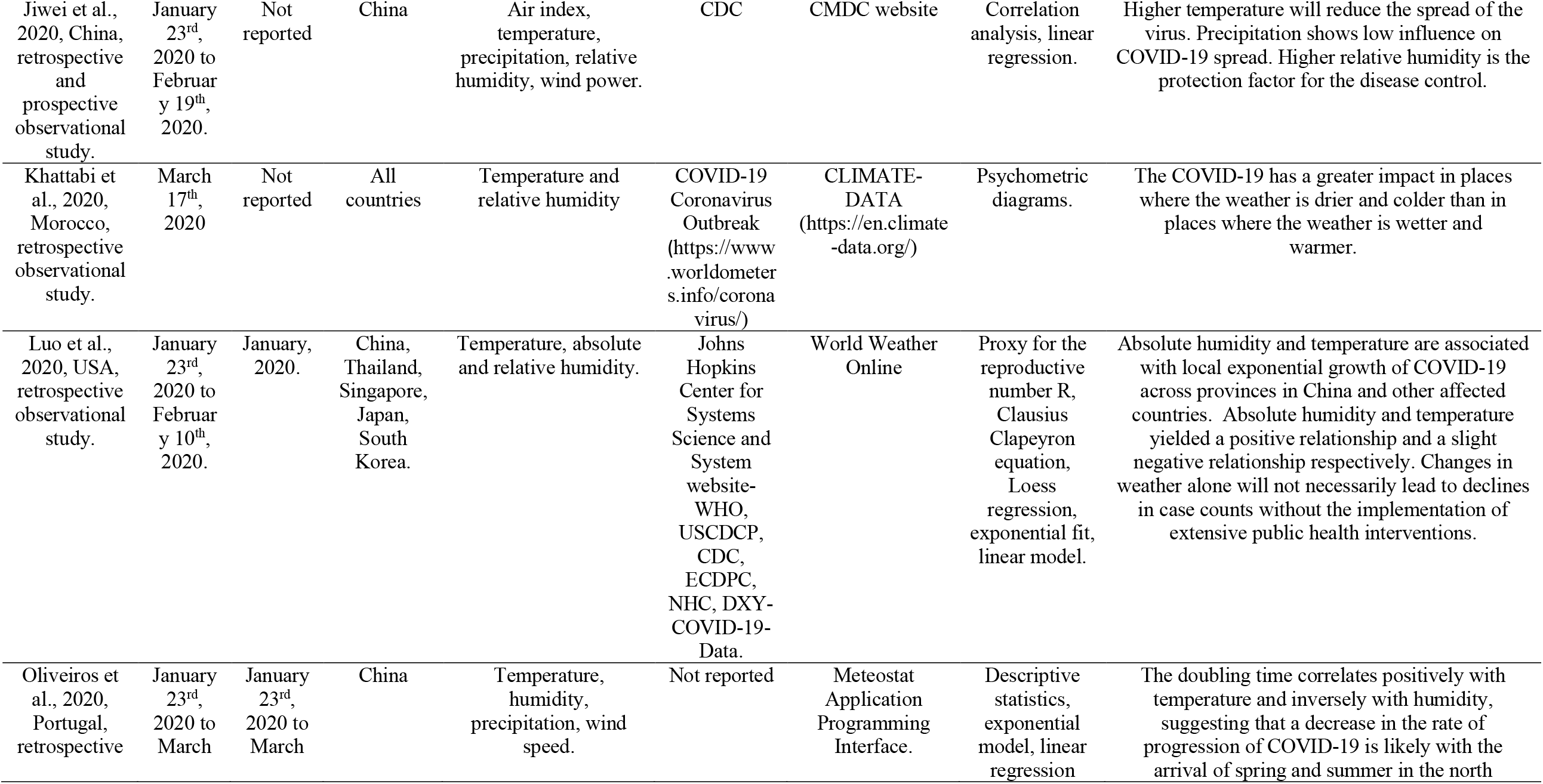
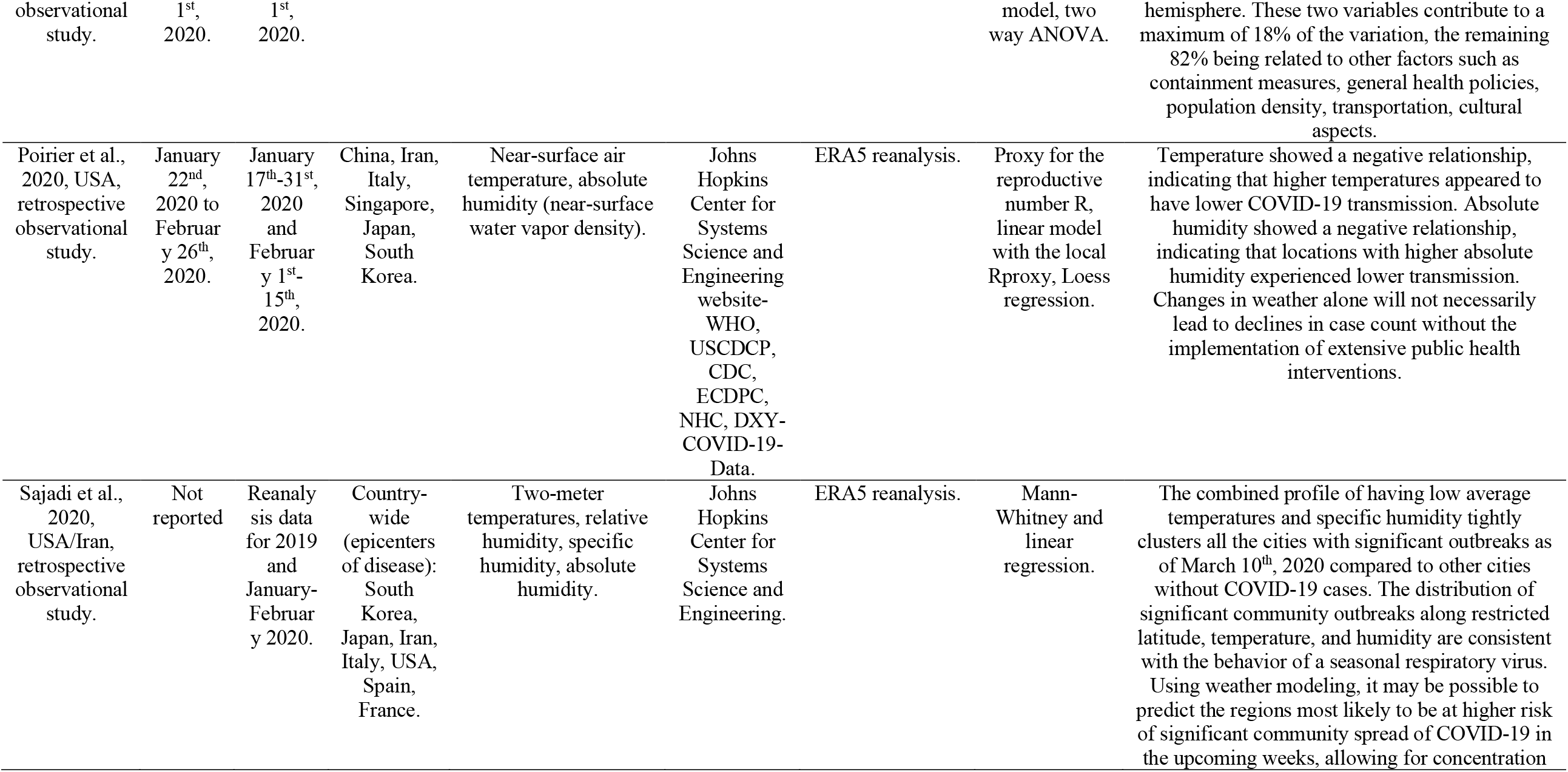
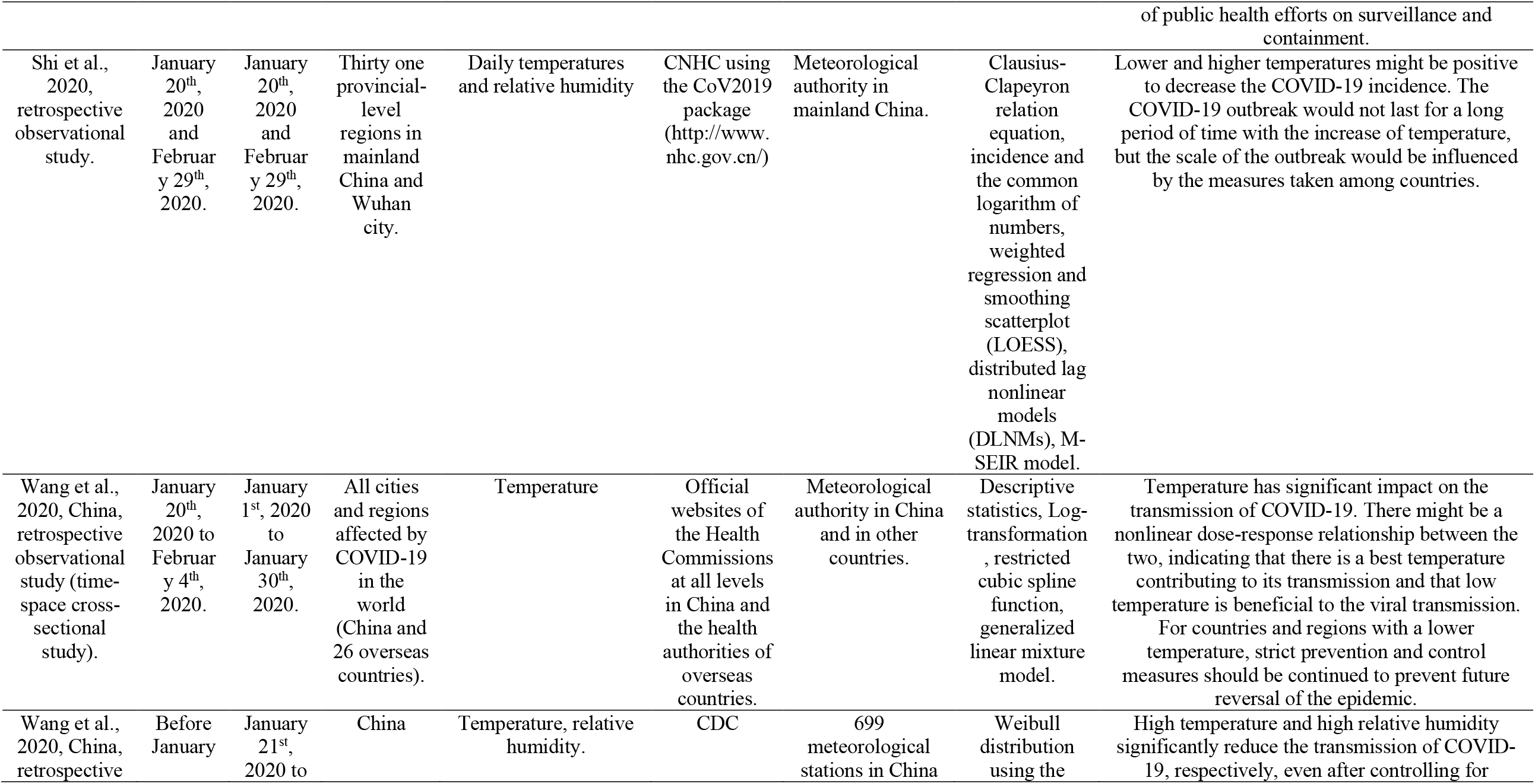
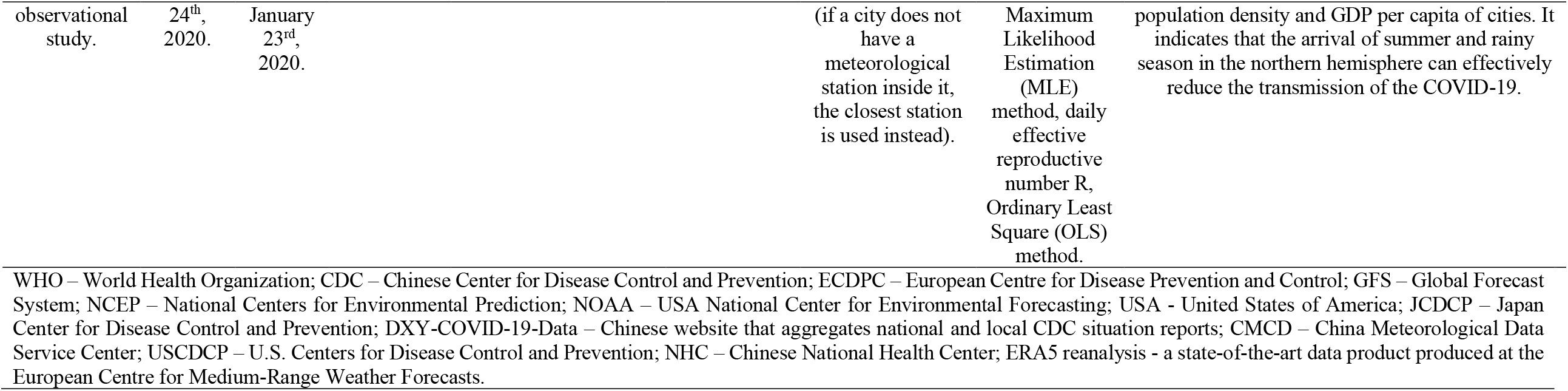
Summary of the data from the studies included in this review.

Great heterogeneity was observed in relation to the displayed variables, that included other weather conditions beyond temperature and humidity[9,20-31] like wind speed,[20,32] visibility,[24] precipitation,[25,27] pressure,[20] rainfall rate,[20] snowfall rate,[20] snow depth,[20] surface downward short-wave irradiation.[20] A heterogeneity was also observed regarding the date of data collection, both in relation to the location studied, and the date of epidemiological data collection of COVID-19 and climatic conditions.

### Results of Individual Studies

Great homogeneity was observed in the results of the effect of temperature and humidity in the seasonal variability and spread of the virus. Sixteen articles selected for final analysis[9,20-30,33-35] were unanimous in stating that cool and dry conditions were potentiating factors for the spread of COVID-19, with the spread being largely absent under extremely cold and very hot and wet conditions. Only one article reported no strong effect of temperature and humidity in the spread of the virus.[32]

It was also noticed that several meteorological factors combined could better describe the epidemic trend than when a single variable was analyzed.[21,24] In addition, confounding factors as public health policies on surveillance and containment, social isolation campaigns (home quarantine strategy), including with patients’ families, socio-economic development contributes to controlling the spread of the virus around the world.[9,20,23-29,31,33,35] Moreover, controlling population density (less crowded cities)[30] and movement,[24] travel limitations,[26,28] increasing the number of medical staff and hospitals, isolating all the suspected cases, understanding the method of each patient’s infection, combining the medical history of the patients with current diagnosis to extract information about the virus,[20] are important measures to combat the new coronavirus.

It was verified, in countries with virus transmission under control like Korea, that the widespread testing to identify potential COVID-19 positive subjects, including asymptomatic ones, could reduce transmission.[23]

Regarding the review process, ten of the selected articles have not yet gone through the peer review process. This must be taken into consideration when making any inference from their conclusion.[9,20-22,24,26-28,31,35]

After considering all these factors, we can infer that confounding variables play an important role, even more significant than temperature and relative humidity, in the spread of COVID-19.

### Synthesis of Results

A meta-analysis was not performed due to the heterogeneity of the methods, locations, and information provided in the related articles investigating the proposed objectives. Additionally, differing units of measure, variables and statistical methods did not allow meaningful comparisons. Only simple and descriptive comparisons were reported, beyond the risk of bias and narrative grade of evidence of the results.

### Risk of Bias Assessment

The risk of bias ranged from high in four papers[9,22,27,32] to moderate in the remaining thirteen (Table 4).[20,21,23-26,28-31,33-35] Limitations were observed in the main items evaluated. Inability to satisfy “the criteria for inclusion in the sample” and “the study subjects and the setting described” items resulted in the low grade, since a limitation in the sample selection criteria meant that none of the studies received a positive evaluation. In addition, many “confounding factors” like public health interventions by the government can interfere with the real effect of the variables studied - temperature and humidity - on the spread of the virus. These factors were only identified in five of the thirteen selected articles.[25,27,28,31,33] In contrast, the items “the exposure measured”, “the outcomes measured” and “the objective, the standard criteria used for measurement of the condition” were, in most articles, performed in a valid and reliable way, except for Oliveiros[27] in all of these three items, Bhattacharjee[32] and Araújo[9] in item “the outcomes measured”, Al-Rousan[20] and Bannister-Tyrrell[21] regarding item “the objective, the standard criteria used for measurement of the condition”.

**Table 4.**
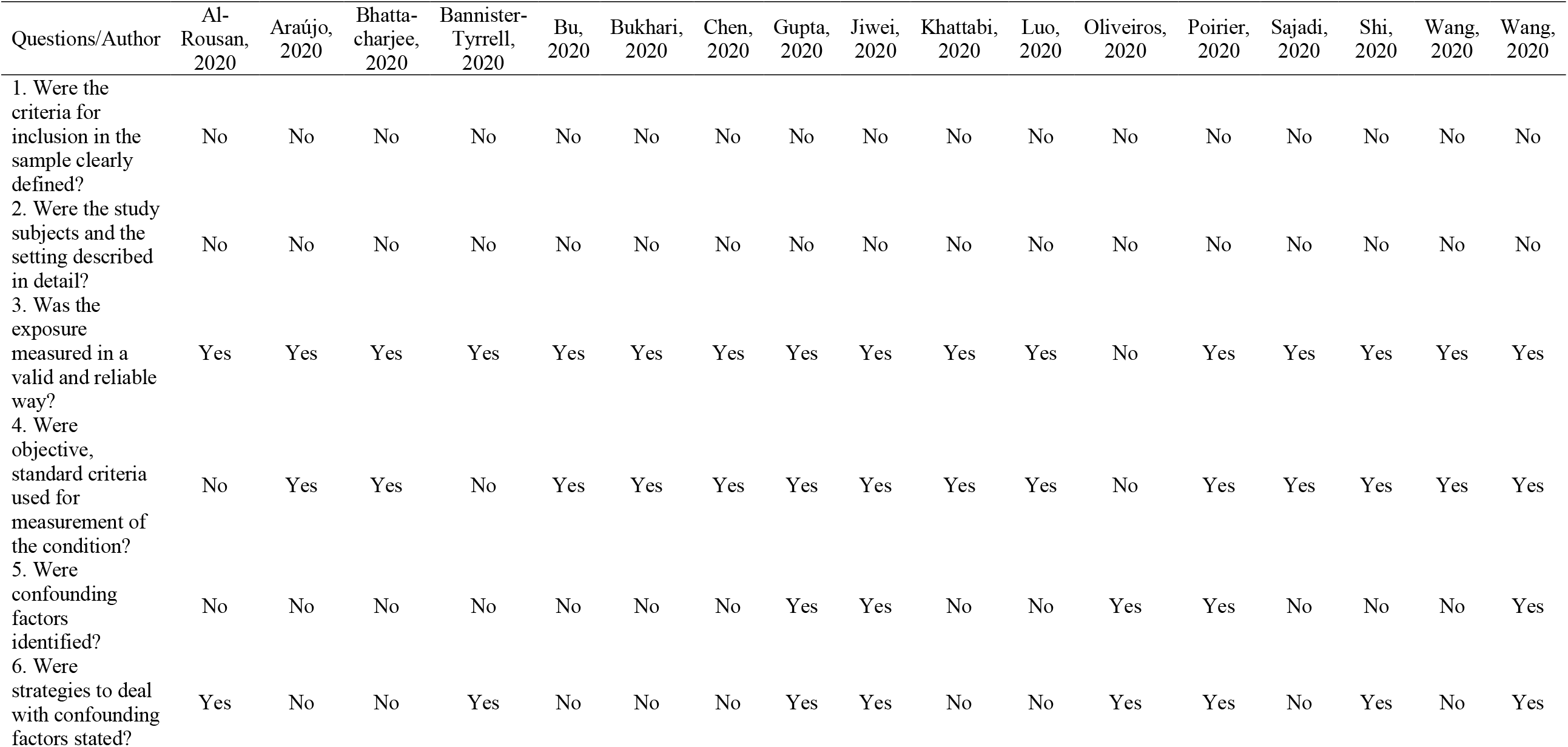
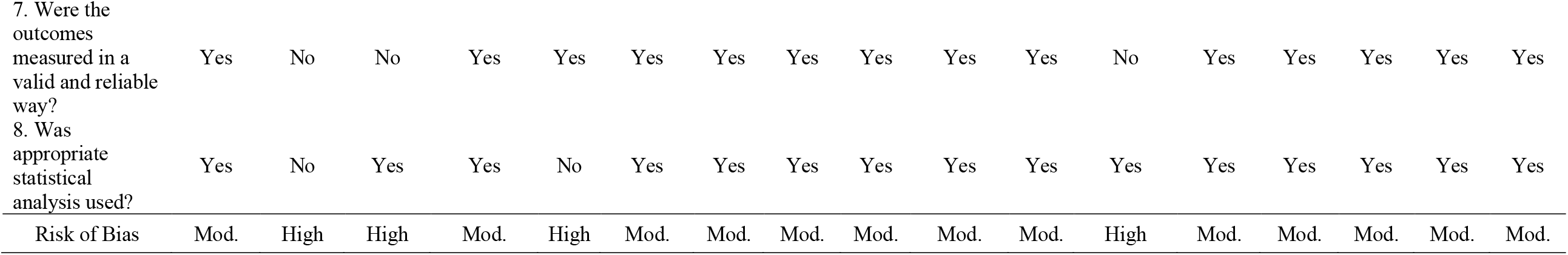
Risk of bias assessment of the studies included in the review.

### Level of Evidence

The evaluation of the certainty of the evidence according to GRADE is described in Table 5. The level of certainty of outcomes evaluated in this systematic review – “Association between temperature and spread rate of COVID-19” and “Association between humidity and spread rate of COVID-19” – were classified as “low”. Since the studies are observational and presented a considerable risk of bias, the certainty of the evidence generated received this classification. Moreover, one study did not show a significant effect of the variables under study on the spread of the new coronavirus.[32]

**Table 5.** Narrative GRADE evidence profile table.

| Outcomes | Impact | N° of Studies | Certainty of the evidence (GRADE) |
| --- | --- | --- | --- |
| Association between temperature and spread rate of COVID-19 | Of the seventeen articles evaluated, sixteen showed some effect of temperature on the transmission rate. Except for one, which concluded that temperature has no effect on SARS-CoV-2 transmission, the other sixteen found that warmer climates are less likely to spread the virus. When other variables are included in the analysis, such as migration patterns, public isolation policies, population density, and cultural aspects, the temperature seems to have less impact. | (17<br>OBSERVATIONAL<br>STUDIES) | ⊕⊕○○<br>LOW<sup>a</sup> |
| Association between humidity and spread rate of COVID-19 | Fourteen manuscripts that investigated the effect of humidity on the transmission of SARS-CoV-2 demonstrated an association between variables. Only one article reported no effect of humidity on the spread of the virus, while the other fourteen showed that wetter climates inhibit the virus spread. As with the temperature, the adjustment for confounding factors decreases the impact of humidity on the transmission of COVID-19. | (15<br>OBSERVATIONAL<br>STUDIES) | ⊕⊕○○<br>LOW<sup>a</sup> |

a. The studies used secondary data obtained through websites for both the number of infected and the climatic conditions. Therefore, eligibility criteria to control possible confounding factors have not been adopted. Furthermore, several studies did not consider variables such as migration patterns and isolation policies in their results, factors that directly impact on the spread rate of SARS-CoV-2/COVID-19.

## Discussion

The results of the articles included in this systematic review indicate that the spread of COVID-19 may be influenced by climatic variables such as temperature and humidity. Apparently, warmer and humid climates may show less transmission of the SARS-CoV-2 virus. Although the level of evidence generated was low, due to the observational design of the studies and the inherent risk of bias, overall great homogeneity of the results was observed among the included studies.

Furthermore, the spread of types of diseases caused by betacoronavirus, such as SARS-CoV-1[11] and MERS-CoV,[36] have already been shown to suffer the impact of climatic conditions. In both these coronoviruses, hot and humid climates demonstrated the ability to decrease the viability of these viruses, while in places with low temperature and humidity there was greater viral stability.

### Summary of evidence

Seventeen articles were included in this systematic review, and methodological issues were identified. Regarding the classification of the articles and their score evaluated by the JBI Critical Appraisal Checklist tool,[18] thirteen studies were classified as moderate risk of bias,[20,21,23-26,28-31,33-35] and four as high risk of bias.[9,22,27,32] This was due, among other factors, to limitations in the sample selection criteria, once the main items “the criteria for inclusion in the sample” and “the study subjects and the setting described” received a negative rating in all selected articles. A reasonable explanation for this fact would be that due to the urgency of the world situation with the rapid spread of COVID-19 worldwide and the need to search for immediate responses to contain the pandemic, secondary data available was used at the moment that the studies were realized, both in relation to the epidemiology of the new coronavirus, and to the climatic conditions, in an attempt to verify a possible association between them. Because of this, there was a limitation in the sample selection criteria and the confounding factors that could interfere with the analysis of these secondary data were not controlled by the eligibility criteria.

The quality of evidence of clinical outcomes was also graded using the GRADE tool. The evidence was scored as low because of the study designs, classified as observational cross-sectional studies, due to the effect of other confounding variables on temperature and humidity in the spread of the virus, and because one selected article did not find a positive strong association between the spread of the new coronavirus with temperature and humidity. According to the author, the results found no strong relationship between the effectiveness of virus and different environmental factors.[32] However, due to homogeneity in the results, which indicate, in summary, that cool and dry conditions were potential factors to the spread of COVID-19, and warmer and wetter climates are less likely to enhance the transmissibility of the virus, the level of evidence could not be rated lower than that.

The analyses of COVID-19 outbreaks in relation to meteorology aspects reveal significant correlations between the incidence of positive cases and climatic conditions. Social factors in combination with meteorological factors play a role in coronavirus outbreaks.[37,38] In fifteen included studies,[9,20,22-30,32-35] the authors investigate the association between temperature and humidity in the transmission rate of COVID-19. In the other two articles,[21,31] the association was made only with temperature.

Luo et al.[26] suggested that sustained transmission and rapid (exponential) growth of cases are possible over a range of humidity and temperature conditions. Bu et al.[22] concluded that a temperature range of 13∼19°C and humidity of 50% ∼ 80% are suitable for the survival and transmission of COVID-19. Moreover, Wang et al.[31] have support the role that temperature could have in changing the COVID-19 human-to-human transmission and that there might be an optimal temperature for the viral transmission. They suggested that colder regions in the world should adopt the strictest social control measures, since low temperatures significantly contribute to the viability, transmission rate and survival of coronaviruses. Finally, Kathabbi et al.[34] stated that by air quality analysis in areas highly contaminated by the virus, the population could be informed and be encouraged to avoid this area, creating a microclimate that helps to eliminate the spread of COVID-19.

Araújo et al.[9] made it clear that it is not possible to characterize the exact local temperature and humidity conditions that minimizing the virus spread. On the contrary, it is reasonable to determine the type of macroclimate conditions in the places where transmission is occurring. For example, in the tropics, where high temperatures and humidity characterize the weather, the climatic suitability for spread of COVID-19 seems to be more difficult. Heat intolerance of the virus is probably related to the breakdown of their lipid bilayer,[39] in a similar model to what occurs with the predecessor of the new coronavirus, the SARS-CoV.[40] Speculative explanation also based on patterns observed for other SARS-CoV justifies the effect of humidity on the less effective spread of the virus on the environment, which reduces the total indirect and secondary transmission.[41] Although higher humidity may increase the atmospheric suspended matter,[22] the amount of virus deposited on surfaces, and virus survival time in droplets on surfaces,[41] the reduction of the virus spread by indirect air transmission may be an important factor behind the reduced spreading of COVID-19 in a humid climate.

High temperature and high humidity reduce the transmission of others infections of the respiratory tract, like influenza[42,43] and of SARS coronavirus.[11,38] The main reasons are: the virus is more stable in cold temperatures, and respiratory droplets, as containers of viruses, remain in suspension longer in dry air.[44] Cold and dry weather can also demote the hosts’ immunity and make them more susceptible to the virus.[45]

Many respiratory pathogens show seasonality and the human activity patterns and immunity can be influenced by environmental factors limited during the COVID-19 outbreak, due to the absence of extreme climatic conditions and specific immunity for a newly emerging virus.[35] Cold air temperature contributes to spread of viruses, including coronavirus, and the possibility of infection. Some possible reasons are: low temperature provides suitable survival and reproduction conditions for coronavirus;[46] cold air causes vasoconstriction of the respiratory tract which contributes to weakening of the immune system; and dry cold air makes the nasal mucosa prone to small ruptures, thereby creating opportunities for virus invasion.[47] In contrast, a long period of low or extremely cold temperature played a positive role in reducing the transmission of the coronavirus.[9,22]

In addition, Bukhari et al.[23] discuss another hypothesis for the lower number of COVID-19 cases detected in the tropics. It could be due to less mass testing as many of the underdeveloped countries that presents deficiency in the health care system and may have not done enough testing to detect the actual spread of this virus.

Two of the articles were classified as retrospective and prospective [24,25] since they suggest implementing future public policies and mass actions that aim to control the spread of COVID-19 around the world. Chen et al.[24] proposed a daily predictive model that in combination with weather observations in the previous 14 days, for five high-latitude cities (New York, Toronto, Italy, Paris and Cologne), is able to predict daily new case counts of COVID-19 for the following 12 days in these places. It is important to notice that a single weather factor alone could not affect the virus transmission too much. However the combination of different meteorological variables could fit a more complex model, in order to address the systematic influence of different types of weather data on the spread of the virus. Jiwei et al.[25] verified further control with specific medicines and an effective vaccine. Considering the practice of social isolation, the process of this strategy is considered a short-time vaccine for susceptible populations in helping to control the disease.

According to Oliveiros et al.,[27] temperature and humidity contribute to a maximum of 18% of the variation, the remaining 82% being related to other factors such as containment measures, general health policies, population density, transportation, and cultural aspects. Population migration is another key factor in the spread process that cannot be ignored,[25] as well as community structure, social dynamics, and global connectivity.[23] In cities with higher levels of population density, the virus is expected to spread faster than that in less crowded cities.[30]

Al-Rousan et al.[20] suggested that international governments should adopt rigorous public policies. This can be done by increasing the number of health professionals and hospitals, and socially isolating mainly the suspected cases. Better health care facilities tend to reduce the transmission of COVID-19.[30] The relatively fast outbreak, associated with imperfect daily reporting practices, make a vast underreporting of new cases of COVID-19. Travel limitations and other control interventions need to be implemented consistently.[28] Additionally, Gupta[33] recommended that all citizens be required to wear a face mask whenever they go out, because the primarily viral infection is through airborne or close contact. The Chinese government used this as a key tool in managing the disease, especially with asymptomatic infected people.

Finally, the review process of the selected articles should be evaluated with caution. Ten of the included studies in this systematic review have not yet gone through the peer review process, which must be taken into consideration when making any inference from the authors’ conclusion.[9,20-22,24,26-28,31,35]

The results of this systematic review indicate that the confounding variables, together, are even more significant than temperature and relative humidity. The timing, implementation, and magnitude of the likely public interventions from international governments should reduce the adverse consequences of COVID-19 on the public health system. Only with proper planning will unnecessary damage be avoided for individuals and the global economy.

### Limitations

The eligibility criteria adopted in the selection of participants were not clear in all included articles. It was only reported that the number of individuals diagnosed with COVID-19 was obtained through secondary data available on websites. In addition, several manuscripts did not go through peer review,[9,20-24,26-28,31,35] due to the urgency of publication on the topic, so care should be taken when considering the results of these studies, although they point in the same direction of effect and impact. Furthermore, some studies[9,20-24,26,29,30] have not evaluated possible confounding factors that could influence the impact of climatic variables on the spread of COVID-19, such as migration patterns, containment measures, general health policies, population density, herd immunity, transportation, and cultural aspects. The articles that included these variables in their analysis[25,27,28,31] concluded that climate alone does not explain most of the variability in the spread of the disease.

## Conclusion

Considering the existing scientific evidence, the spread of COVID-19 seems to be lower in warm and wet climates. However, the certainty of the evidence generated was graded as low. Furthermore, temperature and humidity alone do not explain most of the variability of the COVID-19 outbreak. Public isolation policies, herd immunity, migration patterns, population density, and cultural aspects might directly influence how the spread of this disease occurs.

## Data Availability

Data is available

